# Analysis of the intestinal microbiota in COVID-19 patients and its correlation with the inflammatory factor IL-18 and SARS-CoV-2-specific IgA

**DOI:** 10.1101/2020.08.12.20173781

**Authors:** Wanyin Tao, Guorong Zhang, Xiaofang Wang, Meng Guo, Weihong Zeng, Zihao Xu, Lianxin Liu, Kaiguang Zhang, Yucai Wang, Xiaoling Ma, Zhengxu Chen, Tengchuan Jin, Jianping Weng, Shu Zhu

## Abstract

The current global COVID-19 pandemic is caused by beta coronavirus Severe Acute Respiratory Syndrome Coronavirus-2 (SARS-CoV-2), which already infected over 10 million and caused 500 thousand deaths by June 2020. Overproduction of cytokines triggered by COVID-19 infection, known as "cytokine storm", is a highly risk factor associated with disease severity. However, how COVID-19 infection induce cytokine storm is still largely unknown. Accumulating in vitro and in vivo evidence suggests that gut is also susceptible to COVID19 infection: Human intestinal organoids, an in vitro model which mimic the specific cell type and spatial structure of the intestine, were susceptible to SARS-CoV2 infection; A significant fraction of patients reported gut symptoms; Viral RNA may persist for more than 30 days and infectious virus could be isolated in fecal samples. The gastrointestinal tract is the primary site of interaction between the host immune system with symbiotic and pathogenic microorganisms. The bacteria resident in our gastrointestinal tract, known as gut microbiota, is important to maintain the homeostasis of our immune system. While imbalance of gut microbiota, or dysbiosis, is associated with multiple inflammation diseases5. It's possible that SARS-CoV-2 infection may lead to alternation of gut microbiota thus worsen the host symptom. IL-18 is a proinflammatory cytokine produced multiple enteric cells, including intestinal epithelial cells (IECs), immune cells as well as enteric nervous system, and was shown to increase in the serum of COVID-19 patients. Immunoglobin A (IgA) is mainly produced in the mucosal surfaces, in humans 40-60mg kg-1 day-1 than all other immunoglobulin isotypes combined, and at least 80% of all plasma cells are located in the intestinal lamina propria. Recent study showed that SARS-CoV-2 specific IgA in the serum is positively correlate with the disease severity in COVID-19 patients11. Here we investigated the alterations of microbiota in COVID-19 patients, and its correlation with inflammatory factor IL-18 and SARS-CoV2 specific IgA.

---

The ongoing global pandemic of COVID-19 disease is caused by severe acute respiratory syndrome coronavirus 2 (SARS-CoV-2), which has already infected over 20 million people and caused 700 000 deaths worldwide as of August 2020^1^. Cytokine overproduction triggered by SARS-CoV-2 infection, known as a “cytokine storm”, is highly associated with disease severity. However, how SARS-CoV-2 infection induces a cytokine storm remains unclear. Accumulating *in vitro* and *in vivo* evidence suggests that the gut is also susceptible to SARS-CoV-2 infection. For example, human intestinal organoids, an *in vitro* model that mimics intestinal cellular and spatial structures, are susceptible to SARS-CoV-2 infection^2^, while a significant fraction of COVID-19 patients reported experiencing gut symptoms. Additionally, viral RNA may persist for more than 30 days and infectious virus can be isolated from fecal samples^3,4^. The gastrointestinal tract is the primary site of interaction between the host immune system and symbiotic and pathogenic microorganisms. The bacteria resident in the gastrointestinal tract, known as the gut microbiota, are important for the maintenance of immune homeostasis. A gut microbiota imbalance, or dysbiosis, is associated with multiple inflammatory diseases^5^. It is feasible that SARS-CoV-2 infection may alter the composition of the gut microbiota, thereby worsening COVID-19 symptoms. Interleukin (IL)-18 is a proinflammatory cytokine produced by different types of enteric cells, including intestinal epithelial cells, immune cells, and cells of the enteric nervous system^6,7^, and IL-18 levels are upregulated in the serum of COVID-19 patients ^8^. Immunoglobin A (IgA) is mainly produced in mucosal surfaces. Humans produce more IgA (40-60 mg kg^-1^ day^-1^) than all other immunoglobulin isotypes combined, and at least 80% of all plasma cells are located in the intestinal lamina propria^7,8^. A recent study showed that the serum level of SARS-CoV-2-specific IgA is positively correlated with disease severity in COVID-19 patients^9^. Here, we investigated the changes occurring in the gut microbiota of COVID-19 patients, and the correlation between these alterations and the levels of IL-18 and SARS-CoV-2-specific IgA.

We enrolled 62 COVID-19 patients from the First Affiliated Hospital of the University of Science and Technology of China in Hefei, as well as 33 seasonal flu patients and 40 healthy controls from Hefei. Fresh fecal and serum samples from the three groups were immediately stored at −80 °C for further analysis. We used next-generation sequencing of the V4 region of the 16S ribosomal RNA gene to investigate the potential effect of intestinal SARS-CoV-2 infection on the composition of the gut microbiota. The raw sequencing data were first trimmed and quality-filtered to remove adaptor or low-quality sequences, resulting in an average of 28 054 sequences per sample. The sequences were then further processed using a customized pipeline that combined usearch (v8.1)^10^, vsearch (v2.13.03^11^, and QIIME (v1.9.1)^12^. Alpha-diversity analysis showed that the mean species diversity (Chao1 index) was significantly decreased in COVID-19 patients compared with both flu patients and healthy controls (Fig. 1A). Beta diversity values, as measured by weighted UniFrac, and a histogram depicting the relative taxonomic fraction at the genus level in each group, showed that the abundance and composition of fecal bacteria in COVID-19 patients differed from those of both healthy controls and seasonal flu patients (Fig. 1B and C). To investigate which gut bacteria were associated with COVID-19 patients, linear discriminant analysis effect size (LEfSe) was performed in galaxy (http://huttenhower.sph.harvard.edu/galaxy/) to identify biomarkers for each group. The threshold was set at LDA > 2 to filter the features that differed significantly between COVID-19 patients and seasonal flu patients or healthy controls. Compared with healthy controls, the abundance of members of the genera *Streptococcus, Clostridium, Lactobacillus*, and *Bifidobacterium* was increased, whereas that of members of the genera *Bacteroidetes, Roseburia, Faecalibacterium, Coprococcus*, and *Parabacteroides* was decreased, in SARS-CoV-2-infected patients (Fig. 1D and E). Compared with seasonal flu patients, the abundance of members of the genera *Streptococcus, Veillonella, Fusobacterium, Clostridium, Bifidobacterium*, and *Escherichia* was increased, whereas that of members of the genera *Parabacteroides* and *Sutterella* was decreased, in SARS-CoV-2 patients (Fig. 1F and 1G). The increased abundance of *Streptococcus* in COVID-19 patients was indicative of the risk of infection by opportunistic pathogenic bacteria in this group^13^.

**Figure 1.**
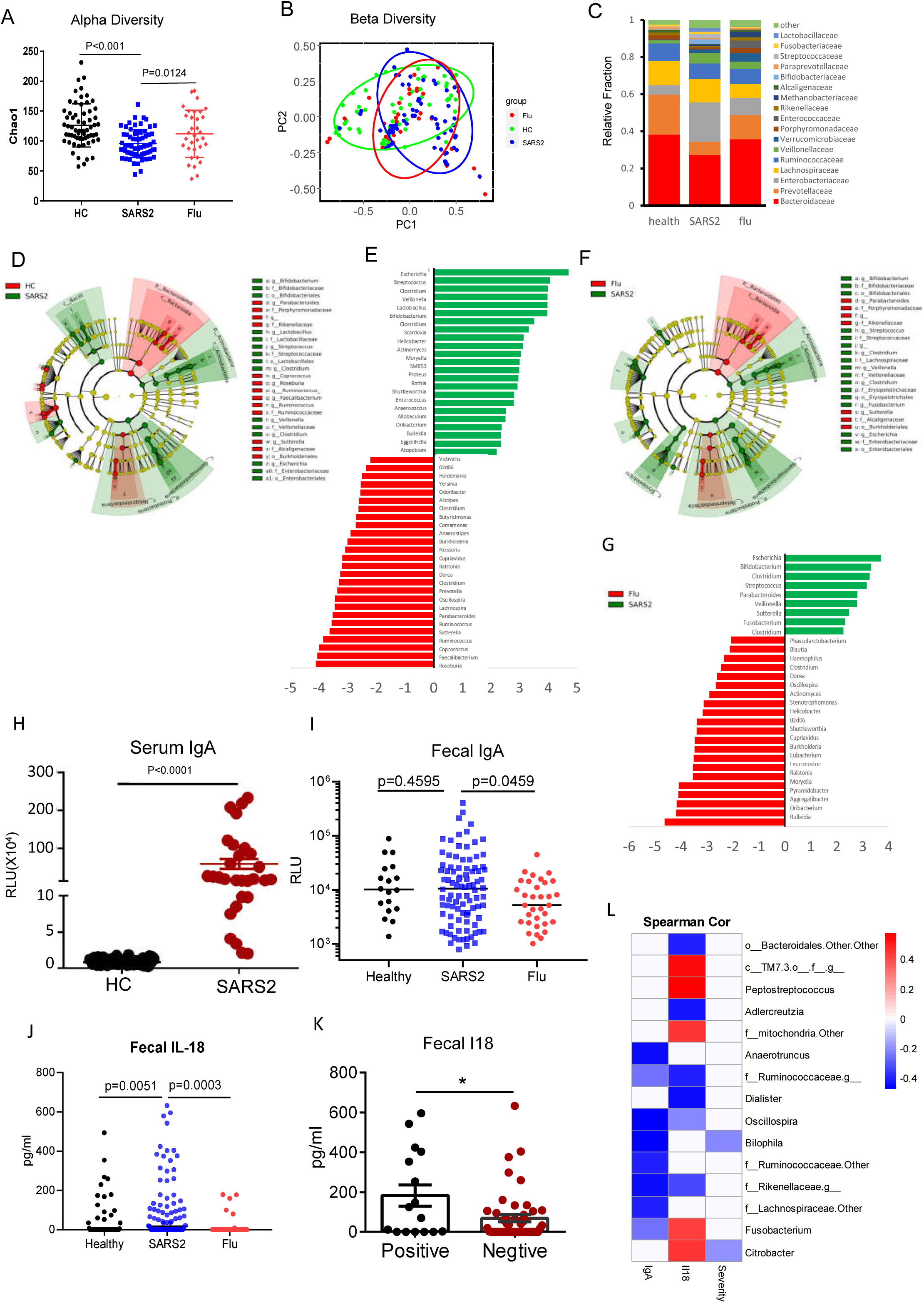
Changes in the gut microbiota of COVID-19 patients. Fecal samples from healthy controls (HC), severe acute respiratory syndrome coronavirus 2 patients (SARS2), and seasonal flu patients (Flu) were first heated at 65 °C to inactivate potential live viruses. Bacterial genomic DNA was extracted from 0.2 g of heat-inactivated fecal samples, amplified using barcoded primers targeting the V4 region of the 16S ribosomal RNA gene, and sequenced on an Illumina MiSeq (paired-end read length of 250 bp). The raw sequencing data were trimmed and processed using a customized pipeline that combined vsearch (v2.13.3), usearch (v8.1), and QIIME (v1.9.1) for analysis of alpha diversity (Chao1) (A) and weighted Unifrac beta diversity (B) of fecal bacteria, as well as to determine the taxonomic fraction in each group (C). Linear discriminant analysis (LDA) effect size (LEfSe) was performed in galaxy to identify biomarkers for each group. (D, E) Features differing significantly (LDA >3.5 for Cladogram plot) between COVID-19 patients and HC. (F, G) Features differing significantly (LDA >3.5 for Cladogram plot) between COVID-19 patients and seasonal flu patients. Fecal samples (0.5 g) were then suspended in 3 ml of sterile PBS buffer and the supernatants used for further analysis. Relative SARS-CoV-2 spike protein-specific immunoglobin A (IgA) antibody concentrations in COVID-19 and flu patients were measured in heat inactivated serum (H) or fecal supernatant (I). (J) The fecal concentrations of interleukin (IL)-18 were measured using an ELISA kit. (K) The fecal concentrations of interleukin (IL)-18 from fecal viral RNA positive or negative COVID-19 patients. (L)The correlation among microbiota composition, gut IL18 concentration, spike protein-specific IgA levels and disease severity were analyzed using Spearman’s correlation coefficient.

Viral intestinal infections induce the production of immunoglobin A (IgA)^8^. Serum samples (0.5 ml) and fecal samples (0.5 g) were both heat-inactivated at 65 °C to prevent potential infection. The fecal samples were then suspended in 3 ml of sterile PBS buffer, and the fecal supernatants together with the serum samples were used for ELISA measurements of SARS-CoV-2-specific IgA. The concentration of spike protein-specific IgA was considerably higher in COVID-19 patients than in either healthy controls (Fig. 1H), suggestive of SARS-CoV-2 mucosal infection in COVID-19 patients. However, there was no significant difference in the concentration of SARS-CoV-2-specific IgA among the fecal supernatants derived from healthy controls, seasonal flu patients, and COVID-19 patients (Fig. 1I). These results indicated that the increased production of SARS-CoV-2-specific IgA was likely due to lung mucosal infection rather than intestinal mucosal infection.

Intestinal infection by viruses can also induce the production of the proinflammatory factor IL-18^14^. Here, we used ELISA to measure the concentrations of IL-18 in the serum samples and fecal supernatants of the three groups. Notably, IL-18 production was higher in both the serum samples and fecal supernatants of COVID-19 patients than in those of either healthy controls or seasonal flu patients (Fig. 1J). Moreover, the IL-18 levels were higher in the fecal supernatants obtained from COVID-19 patients that tested positive for SARS-CoV-2 RNA than those that tested negative (Fig. 1K), indicating that SARS-CoV-2 induced strong intestinal inflammation. We next analyzed the correlation among fecal microbiota composition, Fecal Spike protein specific IgA levels, Fecal IL-18 levels, and disease severity in COVID-19 patients. Intriguingly, fecal IL-18 levels positively correlated with with the relative abundance *of Peptostreptococcus, Fusobacterium* and *Citrobacter* at the genus level (Fig. 1L), indicating that changes in gut microbiota composition might contribute to SARS-CoV-2-induced production of inflammatory cytokines in the intestine and potentially also to the onset of a cytokine storm.

In conclusion, we found SARS-CoV-2-dependent changes in the composition of the gut microbiota of COVID-19 patients. Moreover, higher relative levels of *Streptococcus, Clostridium, Lactobacillus*, and *Bifidobacterium*, and lower relative levels of *Bacteroidetes, Roseburia, Faecalibacterium, Coprococcus*, and *Parabacteroides*, were found in COVID-19 patients when compared with both seasonal flu patients and healthy controls. Interestingly, the concentration of IL-18, a key proinflammatory factor produced by gut cells, was increased in COVID-19 patients, but not in seasonal flu patients. Additionally, fecal supernatant IL-18 levels were higher in COVID-19 patients that tested positive for SARS-CoV-2 RNA than in those that tested negative. Our results suggested that gut microbiota dysbiosis due to SARS-CoV-2 infection may contribute to disease severity, and that IL-18 might serve as an indicator of intestinal infection in COVID-19 patients. Notably, SARS-CoV-2-infected patients were under medical care when their fecal samples were collected, and further studies with treatment-naïve patients are needed to exclude the possibility that changes in the gut microbiota were caused by the administered drugs.

## Data Availability

All data are available upon request.

## Acknowledgements

This work was supported by a grant from the National Key R&D Program of China (2018YFA0508000) (SZ), the Strategic Priority Research Program of the Chinese Academy of Sciences (XDB29030101) (SZ), the National Natural Science Foundation of China (81822021, 91842105, 31770990, 81821001) (SZ), and the Fundamental Research Funds for the Central Universities (WK2070000159) (SZ).

## Author Contribution

WT and SZ designed the experiments. WT, XW, and GZ performed the experiments and interpreted the data. LL, JW, YW, and XM collected all the clinical samples; JW, XM, and KZ provided critical comments and suggestions; TJ, HM, and DZ performed the serological analyses; SZ, WT, and MG wrote the manuscript; SZ and TJ supervised the project.

## Conflict of Interest

We declare no competing interests.

## Notes

### Competing Interest Statement

The authors have declared no competing interest.

### Author Declarations

This is an observational cohort study. This study is part of the project of "Construction of a bio-information platform for novel coronavirus pneumonia (COVID-19) patients follow-up in Anhui" (ChiCTR2000030331). This study was approved by the institutional board of the First Affiliated Hospital of University of Science and Technology of China (2020-XG(H)-009).

## Reference

1 Dodd, D. et al. A gut bacterial pathway metabolizes aromatic amino acids into nine circulating metabolites. Nature 551, 648–652, doi:10.1038/nature24661 (2017).

2 Lamers, M. M. et al. SARS-CoV-2 productively infects human gut enterocytes. Science (New York, N.Y.), doi:10.1126/science.abc1669 (2020).

3 Wu, Y. et al. Prolonged presence of SARS-CoV-2 viral RNA in faecal samples. The lancet. Gastroenterology & hepatology, doi:10.1016/s2468-1253(20)30083-2 (2020).

4 Xiao, F. et al. Infectious SARS-CoV-2 in Feces of Patient with Severe COVID-19. Emerging infectious diseases 26, doi:10.3201/eid2608.200681 (2020).

5 Wu, H. J. & Wu, E. The role of gut microbiota in immune homeostasis and autoimmunity. Gut Microbes 3, 4–14, doi:10.4161/gmic.19320 (2012).

6 Stadnyk, A. W. Intestinal epithelial cells as a source of inflammatory cytokines and chemokines. Canadian journal of gastroenterology = Journal canadien de gastroenterologie 16, 241–246, doi:10.1155/2002/941087 (2002).

7 Fagarasan, S. & Honjo, T. Intestinal IgA synthesis: regulation of front-line body defences. Nature reviews. Immunology 3, 63–72, doi:10.1038/nri982 (2003).

8 Gutzeit, C., Magri, G. & Cerutti, A. Intestinal IgA production and its role in host-microbe interaction. Immunol Rev 260, 76–85, doi:10.1111/imr.12189 (2014).

9 Ma, H. et al. Serum IgA, IgM, and IgG responses in COVID-19. Cell Mol Immunol 17, 773–775, doi:10.1038/s41423-020-0474-z (2020).

10 Edgar, R. C. Search and clustering orders of magnitude faster than BLAST. Bioinformatics 26, 2460–2461, doi:10.1093/bioinformatics/btq461 (2010).

11 Rognes, T., Flouri, T., Nichols, B., Quince, C. & Mahe, F. VSEARCH: a versatile open source tool for metagenomics. PeerJ 4, e2584, doi:10.7717/peerj.2584 (2016).

12 Caporaso, J. G. et al. QIIME allows analysis of high-throughput community sequencing data. Nature methods 7, 335–336, doi:10.1038/nmeth.f.303 (2010).

13 Weiser, J. N., Ferreira, D. M. & Paton, J. C. Streptococcus pneumoniae: transmission, colonization and invasion. Nature reviews. Microbiology 16, 355–367, doi:10.1038/s41579-018-0001-8 (2018).

14 Zhu, S. et al. Nlrp9b inflammasome restricts rotavirus infection in intestinal epithelial cells. Nature 546, 667–670, doi:10.1038/nature22967 (2017).

